# Seroprevalence of SARS-CoV-2 IgG Antibodies in Utsunomiya City, Greater Tokyo, after first pandemic in 2020 (U-CORONA): a household- and population-based study

**DOI:** 10.1101/2020.07.20.20155945

**Authors:** Nobutoshi Nawa, Jin Kuramochi, Shiro Sonoda, Yui Yamaoka, Yoko Nukui, Yasunari Miyazaki, Takeo Fujiwara

**Author notes:** **Corresponding author** Takeo Fujiwara, Department of Global Health Promotion, Tokyo Medical and Dental University (TMDU), 1-5-45 Yushima, Bunkyo-ku, Tokyo 113-8519, Japan.

## Abstract

**Background:** The number of confirmed cases of severe acute respiratory syndrome coronavirus 2 (SARS-CoV-2) infections in Japan are substantially lower in comparison to the US and UK, potentially due to the under-implementation of polymerase chain reaction (PCR) tests. Studies reported that more than half of the SARS-CoV-2 infections are asymptomatic, confirming the importance for conducting seroepidemiological studies. Although the seroepidemiological studies in Japan observed a reported prevalence of 0.10% in Tokyo, 0.17% in Osaka, and 0.03% in Miyagi, sampling bias was not considered. The study objective was to assess the seroprevalence of SARS-CoV-2 in a random sample of households in Utsunomiya City in Tochigi Prefecture, Greater Tokyo, Japan.

**Methods:** We launched the Utsunomiya COVID-19 seROprevalence Neighborhood Association (U-CORONA) Study to assess the seroprevalence of COVID-19 in Utsunomiya City. The survey was conducted between 14 June 2020 and 5 July 2020, in between the first and second wave of the pandemic. Invitations enclosed with a questionnaire were sent to 2,290 people in 1,000 households randomly selected from Utsunomiya basic resident registry. Written informed consent was obtained from all participants. The level of IgG antibodies to SARS-CoV-2 was assessed by chemiluminescence immunoassay analysis.

**Results:** Among 2,290 candidates, 753 returned the questionnaire and 742 received IgG tests (32.4 % participation rate). Of the 742 participants, 86.8% were 18 years or older, 52.6% were women, 71.1% were residing within 10 km from the test clinic, and 89.2% were living with another person. The age and sex distribution, distance to clinic and police district were similar with those of non-participants, while the proportion of single-person households was higher among non-participants than participants (16.2% vs. 10.8%).

We confirmed three positive cases through quantitative antibody testing. No positive cases were found among the people who live in the same household as someone with positive. All cases were afebrile. The estimated unweighted and weighted prevalence of SARS-CoV-2 infection were 0.40% (95% confidence interval: 0.08-1.18%) and 1.23% (95% confidence interval: 0.17-2.28%), respectively.

**Conclusion:** This study suggests the importance of detecting all cases using PCR or antigen testing, not only at a hospital, but also in areas where people assemble. Further prospective studies using this cohort are needed to monitor SARS-CoV-2 antibody levels.

## Introduction

The number of confirmed cases of severe acute respiratory syndrome coronavirus 2 (SARS-CoV-2) infections in Japan are substantially lower in comparison to the US and UK,^1^ potentially due to the under-implementation of polymerase chain reaction (PCR) tests. Studies reported that more than half of the SARS-CoV-2 infections are asymptomatic,^2^ confirming the importance for conducting seroepidemiological studies.^3^ Although the seroepidemiological studies in Japan observed a reported prevalence of 0.10% in Tokyo, 0.17% in Osaka, and 0.03% in Miyagi,^4^ sampling bias was not considered. The study objective was to assess the seroprevalence of SARS-CoV-2 in a random sample of households in Utsunomiya City in Tochigi Prefecture, Greater Tokyo, Japan.

## Methods

We launched the “Utsunomiya COVID-19 seROprevalence Neighborhood Association (U-CORONA)” Study to assess the seroprevalence of COVID-19 in Utsunomiya City. The survey was conducted between 14 June 2020 and 5 July 2020, in between the first and second wave of the pandemic (Figure 1). Invitations enclosed with a questionnaire were sent to 2,290 people in 1,000 households randomly selected from Utsunomiya City’s basic resident registry. Written informed consent was obtained from all participants. The level of IgG antibodies to SARS-CoV-2 was assessed by chemiluminescence immunoassay analysis (Shenzhen YHLO Biotech Co., Ltd., Shenzhen, China).^5^ According to the manufacturer, with a cut-off value of 10 AU/ml, the sensitivity and specificity of the assay was 97.3% and 96.3%, respectively. Using baseline data from the registry, age, sex, distance to clinic, residential district, and the number of cohabitants, multiple imputation was used to estimate the population-based prevalence. All analyses were conducted using STATA 14 (StataCorp LP, College Station, TX, USA) and R version 3.6.0 (R Core Team, 2019). This study was approved by the research ethics committee at Tokyo Medical and Dental University.

**Figure 1.**
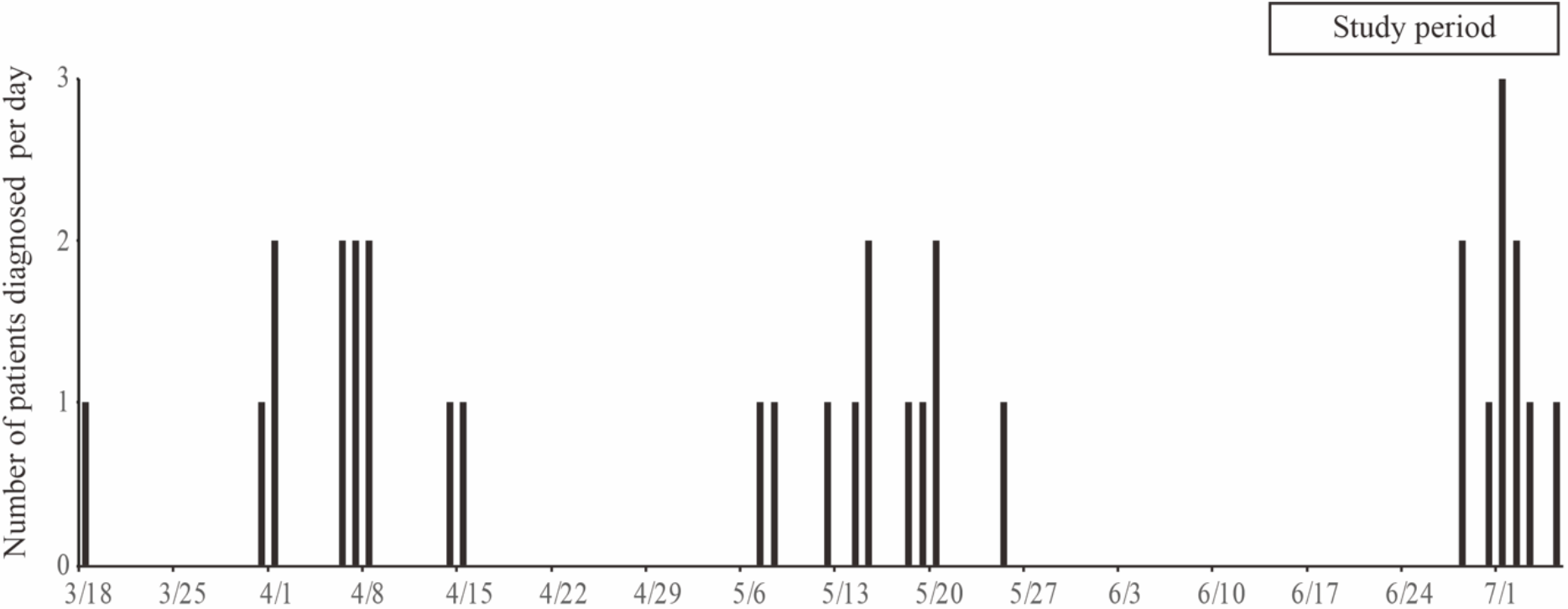
Trend of the number of patients diagnosed with COVID-19 per day between 18 March 2020 and 5 July 2020 in Utsunomiya City, Tochigi Prefecture, Japan (https://www.city.utsunomiya.tochigi.jp/kurashi/kenko/kansensho/etc/1023506.html).

## Results

Among 2,290 candidates, 753 returned the questionnaire and 742 received IgG tests (32.4 % participation rate). Of the 742 participants, 86.8% were 18 years or older, 52.6% were women, 71.1% were residing within 10 km from the test clinic, and 89.2% were living with another person. The age and sex distribution, distance to clinic and police district were similar with those of non-participants, while the proportion of single-person households was higher among non-participants than participants (16.2% vs. 10.8%).

## Discussion

In this household and population-based seroprevalence study in Utsunomiya City, the unweighted and weighted seroprevalence of SARS-CoV-2 were 0.40% and 1.23%, respectively, much lower than Los Angeles in the US (4.31%),^6^ suggesting that the majority of the population in Utsunomiya City was not infected. However, the number of unweighted and weighted estimated cases was 2,074 and 6,378 among 518,610 individuals, respectively, about 90 and 277 times higher than the number of confirmed cases in Utsunomiya City as of 14 June 2020, with 23 cases due to the first pandemic, respectively.

The study limitation is the low participation rate. Also, the study period was relatively long for seroprevalence study. Nonetheless, because we employed a household-based design, participants who do not usually participate in clinical studies may have been pressured to do so by other household members.

This study suggests the importance of detecting all cases using PCR or antigen testing, not only at a hospital, but also in areas where people assemble. Further prospective studies using this cohort are needed to monitor SARS-CoV-2 antibody levels.

## Author Contributions

Dr Fujiwara had full access to all the data in the study and takes responsibility for the integrity of the data and the accuracy of the data analysis.

Concept and design:

Kuramochi, Miyazaki, Fujiwara.

Acquisition, analysis, or interpretation of data:

Nawa, Kuramochi, Sonoda, Yamaoka, Nukui, Miyazaki, Fujiwara

Drafting of the manuscript:

Nawa, Fujiwara.

Critical revision of the manuscript for important intellectual content:

Nawa, Kuramochi, Sonoda, Yamaoka, Nukui, Miyazaki, Fujiwara.

Statistical analysis:

Nawa, Fujiwara.

Administrative, technical, or material support:

Nawa, Kuramochi, Sonoda, Yamaoka, Nukui, Miyazaki, Fujiwara.

Supervision: Fujiwara.

## Data Availability

All data is included in the manuscript.

## Acknowledgement

We thank Yuna Koyama, MD-PhD candidate at TMDU, Hisaaki Nishimura, MD, MSc, PhD candidate, Euma Ishii, MD-PhD candidate, Yoshifumi Fukuya, MD, PhD candidate, Keitaro Miyamura, MD, PhD candidate, and Yu Funakoshi, MD, MPH, PhD candidate, all at TMDU. We also thank medical students at TMDU who participated in the data collection, medical staff in Kuramochi Clinic Interpark, and all the participants in this study.

## Funding

This research was supported by the Japan Agency for Medical Research and Development (AMED) under Grant Number 2033648.

**Table 1.** Demographic characteristics of study participants

|  |  | Participants in unimputed dataset (n=742 from 341 households) | Number and prevalence of positive test in unimputed dataset (n=742) | Estimated prevalence in imputed dataset (n=2,290) |
| --- | --- | --- | --- | --- |
| Variable |  | N or Mean (% or SD) | N (%; 95% CI<sup>b</sup>) | % (95% CI<sup>c</sup>) |
| Whole sample |  | 742 (100) | 3 (0.40, 0.08, 1.18) | 1.23 (0.17, 2.28) |
| Age (in years) | <10 | 56 (7.6) | 0 (0, 0, 6.38) | NA |
|  | 10-17 | 42 (5.7) | 0 (0, 0, 8.41) | NA |
|  | 18-65 | 463 (62.4) | 3 (0.65, 0.13, 1.88) | 1.29 (0.14, 2.43) |
|  | ≥65 | 181 (24.4) | 0 (0, 0, 2.02) | NA |
| Sex | Male | 352 (47.4) | 3 (0.77, 0.16, 2.23) | 1.90 (0.05, 3.76) |
|  | Female | 390 (52.6) | 0 (0, 0, 1.04) | NA |
| Distance to clinic (km) | <1 | 4 (0.5) | 0 (0, 0, 60.24) | NA |
|  | 1-10 | 524 (70.6) | 2 (0.38, 0.05, 1.37) | 1.04 (0, 2.15) |
|  | ≥10 | 214 (28.8) | 1 (0.47, 0.01, 2.58) | 1.42 (0, 4.07) |
| Residential district | Center | 283 (38.1)<sup>a</sup> | 1 (0.35, 0.009, 1.95) | 1.11 (0, 2.85) |
|  | East | 290 (39.1)<sup>a</sup> | 2 (0.69, 0.08, 2.47) | 1.55 (0, 3.45) |
|  | South | 169 (22.8)<sup>a</sup> | 0 (0, 0, 2.16) | NA |
| Number of cohabitants | 1 | 80 (10.8)<sup>a</sup> | 0 (0, 0, 4.51) | NA |
|  | 2 | 211 (28.4)<sup>a</sup> | 0 (0, 0, 1.73) | NA |
|  | ≥3 | 451 (60.8)<sup>a</sup> | 3 (0.67, 0.14, 1.93) | 1.50 (0.02, 2.98) |

<sup>a</sup> $p < 0.05$ for comparison between those who participated and those who did not. Residential district was significantly different between non-participants and participants (south, 28.0% vs 22.8% and center, 31.6% vs. 38.1%, respectively).
<sup>b</sup> Confidence intervals estimated using exact binomial distribution.
<sup>c</sup> Confidence intervals estimated by logit-transformed standard error for 50 times and combined by Rubin's rules.
NA: not available.

